# The combined impact of persistent infections and human genetic variation on C-reactive protein levels

**DOI:** 10.1101/2022.01.07.22268880

**Authors:** F. Hodel, O. Naret, C. Bonnet, N. Brenner, N. Bender, T. Waterboer, P. Marques-Vidal, P. Vollenweider, J. Fellay

## Abstract

Multiple human pathogens establish chronic, sometimes life-long infections. Even if they are often latent, these infections can trigger some degree of local or systemic immune response, resulting in chronic low-grade inflammation. There remains an incomplete understanding of the potential contribution of both persistent infections and human genetic variation on chronic low-grade inflammation. We searched for potential associations between seropositivity for 13 persistent pathogens and the plasma levels of the inflammatory biomarker C-reactive protein (CRP), using data collected in the context of the UK Biobank and the CoLaus|PsyCoLaus Study, two large population-based cohorts. We performed backward stepwise regression starting with the following potential predictors: serostatus for each pathogen, polygenic risk score for CRP, as well as demographic and clinical factors known to be associated with CRP. We found evidence for an association between *Chlamydia trachomatis* (P-value = 5.04e-3) and *Helicobacter pylori* (P-value = 8.63e-4) seropositivity and higher plasma levels of CRP. We also found an association between pathogen burden and CRP levels (P-value = 4.12e-4). These results improve our understanding of the relationship between persistent infections and chronic inflammation, an important determinant of long-term morbidity in humans.

## Introduction

Inflammation is a complex and necessary response of the immune system to harmful stimuli such as tissue injury, infection, or exposure to toxins (1). During the acute phase that is characterized by blood flow changes and increased blood vessel permeability, plasma proteins and leukocytes migrate from the circulation to the site of inflammation (2). This immediate protective response usually enables the elimination of the initial cause of the cell injury and the restoration of homeostasis. However, when the acute response fails to clear tissue damage, for example because of prolonged exposure to stimuli, inflammation can become a chronic process (3). A number of common diseases are at least partly caused by chronic inflammation, including coronary artery disease, type 2 diabetes, and some cancers (4). Thus, although inflammation plays an important role in human defense against aggression, it also contributes to the pathophysiology of multiple diseases of major public health importance.

Diagnostic tests are capable of detecting the presence and intensity of systemic inflammation (5). The most commonly used inflammatory biomarker is the acute phase reactant Creactive protein (CRP). This ring-shaped protein is produced by hepatocytes upon stimulation by pro-inflammatory cytokines such as interleukin (IL)-1β, IL-6, and TNF-α.

Although CRP is commonly used as a sensitive indicator of inflammation, the factors influencing its baseline plasma levels are only partially understood. Circulating amounts of CRP are positively associated with age, body mass index (BMI) and smoking, and inversely with male sex and physical activity (6–8). In addition, large-scale genomic analyses have shown that common genetic variation explains up to 7.0% of the variance in plasma CRP levels (9).

To get a more comprehensive view of the factors influencing chronic inflammation in the general population, we used samples and data from the UK Biobank and the CoLaus|PsyCoLaus study to search for associations between baseline CRP levels and chronic infection by persistent/latent pathogens, after careful adjustment for all known demographic, clinical and genomic influences. Indeed, some infectious agents causing long-term infections in humans have been shown to trigger some degree of local or systemic immune response, resulting in a chronic state of low-grade inflammation that may lead to deleterious health outcomes (10, 11).

## Methods

### Study cohorts. The UK Biobank

is a population-based exploratory study of which the enrollment procedure has been outlined previously (12). In brief, half a million men and women between the ages of 40 and 69 (45.6% male, mean age ± SD: 56.5 ± 8.1) visited one of 22 UK Biobank screening centers in England, Scotland and Wales between 2006 and 2010. The evaluation included a survey, a personal interview, and a number of physical measurements and blood. Urine and saliva samples were also collected for long-term storage. This research was undertaken with approved access to UK Biobank data under application number 50085 (PI: Fellay). All UK Biobank study participants gave informed consent at the time of recruitment. Ethical approval for the UK Biobank study was obtained from the North West Centre for Research Ethics Committee (11/NW/0382).

### The CoLaus|PsyCoLaus study

is a prospective population-based study initiated in 2003 in Lausanne, Switzerland (www.colaus-psycolaus.ch) (13). It involves more than 6’000 participants of European ancestry (47.5% male) initially aged 35 to 75 years (mean ± SD: 51.1 ± 10.9), thus representing a sample of approximately 10% of the inhabitants of Lausanne. Individuals were randomly recruited from the general population and are monitored every 5 years regarding their lifestyle and health status. Detailed phenotypic information was obtained from each study participant through questionnaires, physical assessment, and biological measurements of blood and urine markers. The institutional Ethics Committee of the University of Lausanne, which afterward became the Ethics Commission of Canton Vaud (www.cervd.ch) approved the baseline CoLaus|PsyCoLaus study (reference 16/03, decisions of 13th January and 10th February 2003), and written consent was obtained from all participants.

### DNA genotyping and quality checks

Genotyping and imputation of UK Biobank individuals have been fully described by Bycroft *et al*. (14). Briefly, samples were genotyped on either the UK BiLEVE Axiom array (Affymetrix) or UK Biobank Axiom array (Applied Biosystems). Genotypes were phased using SHAPEIT3 and the 1000 Genome phase 3 dataset as a reference, then imputed using IMPUTE4 using the Haplotype Reference Consortium data, 1000 Genomes phase 3, and UK10K data as references (15–17). Post-imputation quality checks resulted in a total number of 9’349’624 single nucleotide polymorphisms (SNPs) available for analyses. DNA samples from 5’399 CoLaus|PsyCoLaus participants were genotyped for 799’653 SNPs using the BB2 GSK-customized Affymetrix Axiom Biobank array. Quality control procedures and imputation of genotypes have been previously described in Hodel *et al*. (18). A total of 9’031’263 SNPs from the CoLaus|PsyCoLaus dataset were included for further analyses (flowchart of inclusion/exclusion criteria in Supplementary Fig. 1).

### Measurement of inflammatory biomarkers

For the UK Biobank, non-fasting venous blood samples (∼ 50 mL) were collected at recruitment. Blood samples were shipped at 4°C to the central processing and archiving facility in Stockport. Serum high-sensitivity CRP (hs-CRP) concentrations were measured in participants by immunoturbidimetric assay on a Beckman Coulter AU5800. The manufacturer’s analytical range was 0.08 to 80 mg/L. Ninety-five individuals with a hsCRP level 20 mg/L were removed from the analysis. For CoLaus|PsyCoLaus, venous blood samples (∼ 50 mL) were drawn in the fasting state and allowed to clot. Serum blood samples were kept at 80°C before assessment of cytokines and sent in dry ice to the laboratory. Hs-CRP was assessed by immunoassay and latex HS (IMMULITE 1000–High, Diagnostic Products Corporation, LA, CA, USA). For quality control, repeated measurements were conducted in 80 subjects randomly drawn from the initial sample. Forty-seven individuals with hs-CRP levels above 20 mg/L were assigned a value of 20 by the manufacturer and were therefore removed from the hs-CRP analyses as they are indicative of acute inflammation.

### Serological analyses

To assess humoral responses to a total of 56 antigens derived from 24 persistent infectious agents (45 antigens from 20 pathogens in UK Biobank, and 38 antigens from 18 pathogens in CoLaus|PsyCoLaus), serum samples were independently analyzed by the Infections and Cancer Epidemiology Division at the German Cancer Research Center (Deutsches Krebsforschungszentrum, DKFZ) in Heidelberg (19, 20). Seroreactivity was measured at serum dilution 1:1000 using multiplex serology based on glutathione-S-transferase (GST) fusion capture immunosorbent assays combined with fluorescent bead technology. For each infectious agent tested, antibody responses were measured for one to six antigens and then expressed as a binary result (IgG positive or negative) based on predefined median fluorescence intensity (MFI) thresholds (21). For our analysis, only antigens shared between the two cohorts were retained, resulting in a final combination of 27 antigens from 13 pathogens. To define overall seropositivity against infectious agents when more than one antigen was used, we applied the pathogen-specific algorithms suggested by the manufacturer. Details of the methods on how the antigens were combined have been described previously (21).

### Combining study cohorts

Upon completion of the genotyping and quality control (QC) analyses for each cohort, imputed datasets were matched on strand, SNP ID and genomic coordinates. Additional analyses and QC checkpoints were performed to ensure proper merging. This resulted in a dataset of 12’055 unique individuals of European ancestry and a total of 6’899’629 markers.

### Genome-wide association study of hs-CRP and tissue prioritization

Single-marker genome-wide association study (GWAS) of log-10 transformed hs-CRP was conducted using a general linear model association analysis in PLINK 2.0, adjusting for sex, age, BMI, and the top 10 principal components (PCs) of the genotyping data (22). Some individuals were excluded at this stage due to missing covariate data and GWAS was thus performed on a final set of 12’031 individuals. We performed differentially expressed gene (DEG) analysis using the GENE2FUNC procedure in FUMA (23). FUMA defined the DEGs in each tissue by performing a two-sided Student’s t-test for each tissue against the remaining tissues. Each of the 30 general tissue types available from GTEx v8 was tested for up-regulation, down-regulation, and both-sided DEG sets (24). Genes with a P-value < 0.05 and an absolute fold change *≥* 1.5 (0.58 in log2 scale) were defined as DEG.

### Polygenic risk score calculation for hs-CRP level

We carried out a polygenic risk score (PRS) analysis to investigate the relationship between human genetic variation and hs-CRP levels. A CRP-PRS was calculated for each study participant based on the risk effects of common SNPs derived from GWAS summary statistics of hs-CRP. As a baseline cohort, we referred to the GWAS summary statistics of the CHARGE cohort (N= 204’402, heritability h2 = 6.5%) (9, 25). These summary statistics were used to construct the CRP-PRS in our target cohort consisting of the merged UK Biobank and CoLaus|PsyCoLaus data using the clumping and thresholding method of the PRSice-2 v2.2.7 software (26). We used a standardized method to obtain PRS, by multiplying the dosage of risk alleles for each variant by the effect size in the GWAS and summing the scores across all of the selected variants. SNPs were clumped based on linkage disequilibrium (LD) (r2 *≥* 0.1) within a 250 kb window. Model estimates of the PRS effect were adjusted for sex, age, BMI, and the top 10 PCs. As an additional quality control, the distribution of PRS was checked in each cohort separately, to ensure that they followed a normal distribution.

### Analyses of the determinants of hs-CRP levels

We used linear regression with backward selection to identify the factors significantly associated with hs-CRP plasma levels. Tested covariates included serostatus for each pathogen, polygenic risk score for CRP, age, sex, BMI, and the first 10 PCs of the genotyping data. P-value < 0.05 was considered statistically significant. The analysis was performed using the stepAIC function in R version 4.0.5 (27).

## Results

### Baseline characteristics of study participants

We studied a total of 12’055 individuals with available hs-CRP level, serological results, and genome-wide genotyping data from two independent population-based studies: the UK Biobank (N = 8’371) and the CoLaus|PsyCoLaus study (N = 3’684) (Supplementary Fig. 1). Participants ranged in age from 35 to 75 years (mean age ± SD: 55.68 ± 9.07), with a majority of women (55.4%) and a mean BMI of 26.80 (± SD: 4.73). HsCRP level was measured in all participants. The median hsCRP level was 1.30 mg/L (10th, 90th percentiles: 0.35 mg/L, 5.10 mg/L). Fig. 1 shows the distributions of age, sex, BMI and log10-transformed hs-CRP in both cohorts. We observed a very comparable distribution of all relevant variables in the two cohorts, which were merged for downstream analyses. Supplementary Fig. 2 shows associations of hs-CRP with demographic and clinical factors. Higher levels of hs-CRP associated with female sex, age and increased BMI (P-values = 1.5e-3, 3.4e-69, and ≈ 0, respectively).

**Fig. 1.**
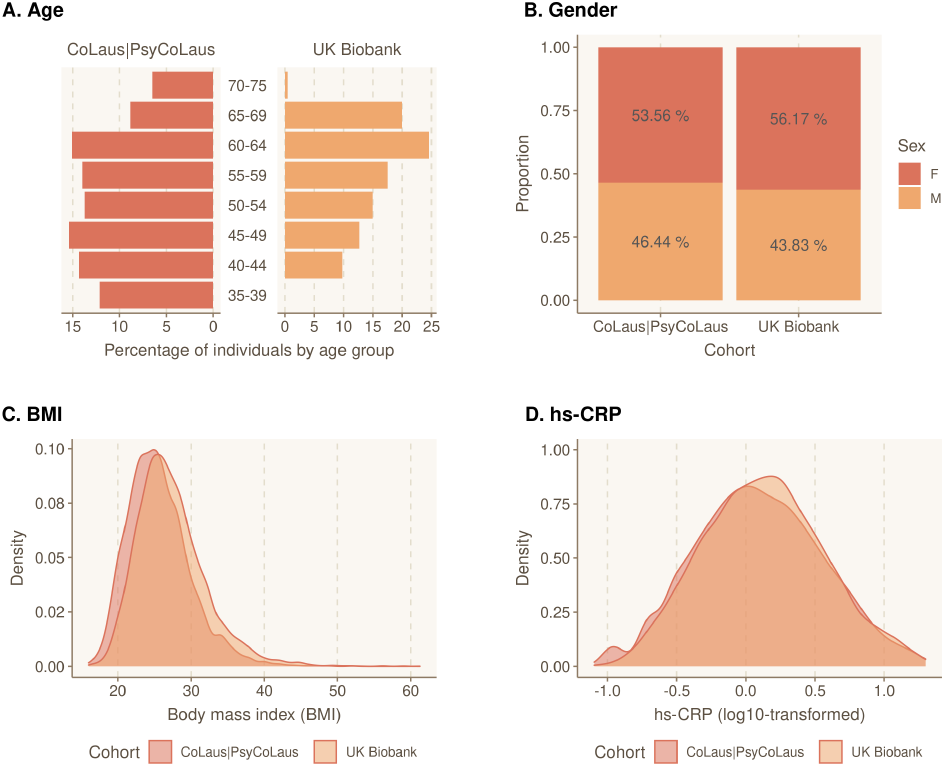
Baseline characteristics of the study cohort. Distribution of A) age, B) gender, C) body mass index (BMI) and D) hs-CRP for participants by subcohort.

### The impact of genetic variation on hs-CRP levels

The filtered genetic variants from the two cohorts were combined (see Methods) to increase sample size. To estimate sample variation, and to control for potential population structure and genotyping bias, principal component analysis (PCA) was performed using the correlation matrix of the genotyping data. PCA plots for the first ten principal components (PC1-PC10) are shown in Supplementary Fig. 3A, annotated by the original cohort from which the sample was drawn. We observed that samples from both subgroups (UK Biobank and CoLaus|PsyCoLaus) were segregated on the first PC (PC1) and eighth PC (PC8), but not on the other PCs. The top 10 PCs explained 61% of the total variance and were used throughout the study to correct for stratification (Supplementary Fig. 3B).

We performed GWAS in our merged data using a continuous, quantitative approach to search for human genetic determinants of hs-CRP levels. Factors associated with hs-CRP levels (age, sex, and BMI) were included as covariates in the linear regression models, in addition to the coordinates of the top 10 PCs (Supplementary Fig. 2). The Manhattan plot for the GWAS results is presented in Fig. 2. Our study replicates six previously reported associated loci involved in immune and hepatic metabolic pathways, including rs4655584 in *LEPR* (P-value = 5.57e-27), rs4129267 in *IL6R* (P-value = 4.23e-19), rs2794520 in *CRP* (P-value = 2.51e-32), rs780093 in *GCKR* (P-value = 1.85e-13), rs7970695 in *HNF1A* (P-value = 2.32e-29) and rs429358 in the *APOE* locus (P-value = 3.30e-51) (9, 28–31). Quantile-quantile (Q-Q) plot and genomic control lambda (l= 1.03) revealed no deviation in the test statistics compared with the expectation (Supplementary Fig. 4). We then performed differential expression analysis (DEA) across GTEx v8 RNA-seq data for 30 general tissue types on the mapped GWAS-associated genes and identified a significant enrichment of upregulated genes in human liver (Supplementary Fig. 5).

**Fig. 2.**
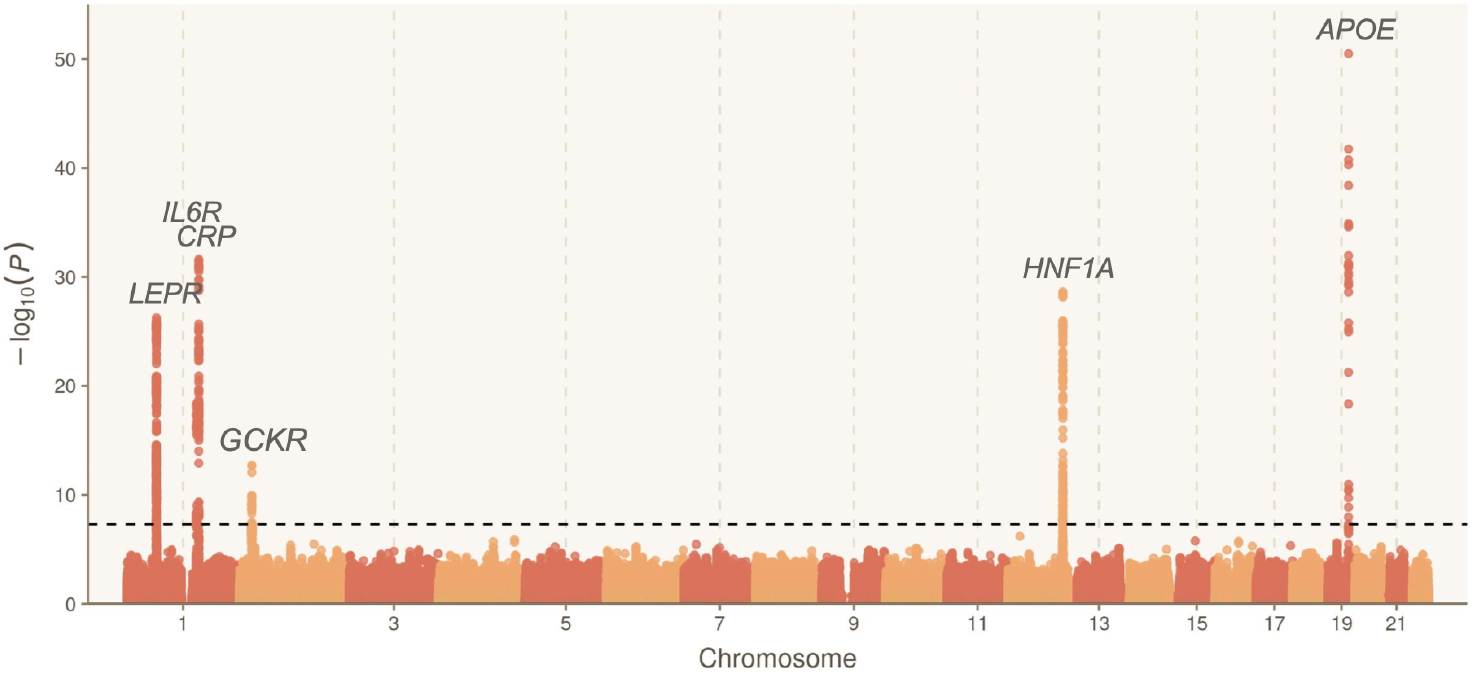
Manhattan plot of genome-wide association results of hs-CRP levels. Manhattan plot showing the significance of association of all SNPs across chromosomes 1 to 22. SNPs are plotted on the x-axis according to their physical position on each chromosome and the strength of the association with serostatus is indicated on the y-axis as -log10 P-value. The dashed line marks the Bonferroni-corrected genome-wide significance threshold of 5e-08. Gene labels are annotated as the nearby genes to the significant SNPs.

Next, we computed a CRP-PRS to investigate the effect of multiple gene variants on hs-CRP levels. A total of 1’809 SNPs were included at the best P-value threshold (P-value = 3.65e-3). The PRS followed a normal distribution in the merged cohort, as well as in each subcohort separately (Supplementary Fig. 6). To describe the influence of common human genetic variation on plasma hs-CRP levels, we quantified the trait variance (R2) explained by the derived PRS and covariates across individuals. We observed that the variance explained by the full model was 25.8%, with 21.5% attributed to the demographic and clinical covariates and 4.3% to the CRP-PRS. The association between the CRP-PRS and hs-CRP levels was very strong (P-value = 6.58e-123; Supplementary Fig. 7), with hs-CRP levels increasing by 0.48 [Standard error (SE) 0.02] for each standard deviation increment in CRP-PRS.

### Associations between persistent/latent infections and hs-CRP levels

We searched for associations between hsCRP levels and serostatus for the following persistent or frequently recurring human pathogens: 10 viruses (BK virus (BKV), Cytomegalovirus (CMV), Epstein–Barr virus (EBV), Human Herpes Virus (HHV)-6, HHV-7, Herpes Simplex Virus (HSV)-1, HSV-2, JC virus (JCV), Kaposi’s sarcoma-associated herpesvirus (KSHV) and Varicella zoster virus (VZV)); two bacteria (*Chlamydia trachomatis (*C. trachomatis) and *Helicobacter pylori* (*H. pylori*)); and one parasite (*Toxoplasma gondii* (*T. gondii*)) (Fig. 3). The overall seropositivity ranged from 6.57% (KSHV) to 95.25% (EBV). Cohort-separated seroprevalences are shown in Supplementary Fig. 8 and 9.

**Fig. 3.**
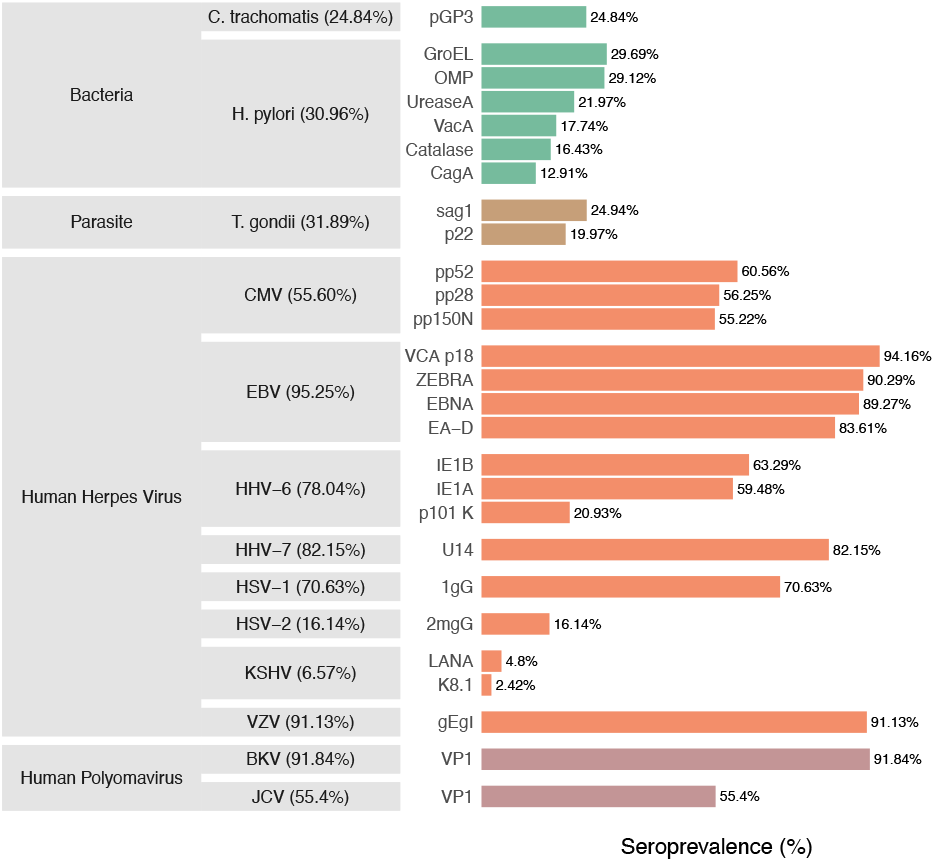
Overall pathogen seropositivity and seroprevalence of tested antigens. List of the 13 pathogens and 27 antigens available from the combined study. The grey boxes indicate the pathogen on which the antigen protein is found, and the family to which the pathogen belongs. Percentages in parentheses after pathogen names indicate overall seropositivity for the specified pathogen. The percentages on the right indicate the seroprevalence of antibodies against infectious disease antigens tested using Multiplex Serology platform. For study-based figure see Supplementary Fig. 8 and 9.

Using backward stepwise regression, we observed associations of hs-CRP levels with seropositivity for *H. pylori* (P-value = 8.63e-4) and *C. trachomatis* (P-value = 5.04e-3) (Table 1). The final regression model including these two pathogens, sex, age, BMI, and the top 10 PCs, explained 25.9% of the variance of hs-CRP levels. This explained fraction of the variance is similar to the value obtained without including the serological results (above), indicating that the impact of *H. pylori* and *C. trachomatis* seropositivity on chronic inflammation, even if statistically significant, is likely to be minimal at the population level.

**Table 1.** Linear regression analysis results for hs-CRP.

| Characteristic | Beta | 95% CI<sup>1</sup> | P-value |
| --- | --- | --- | --- |
| *C. trachomatis* |  |  | 0.005 |
| Seronegative | - | - |  |
| Seropositive | 0.02 | 0.01, 0.04 |  |
| *H. pylori* |  |  | <0.001 |
| Seronegative | - | - |  |
| Seropositive | 0.03 | 0.01, 0.04 |  |
| Age | 0.01 | 0.01, 0.04 | <0.001 |
| BMI | 0.04 | 0.04, 0.04 | <0.001 |
| Sex |  |  | <0.001 |
| M | - | - |  |
| F | 0.07 | 0.06, 0.09 |  |
| CRP-PRS | 0.10 | 0.09, 0.10 | <0.001 |
| PC1 | 2.0 | 1.1, 3.0 | <0.001 |
| PC2 | -1.1 | -1.9, -0.27 | 0.008 |
| PC3 | 2.3 | 1.5, 3.1 | <0.001 |
| PC4 | -1.5 | -2.3, -0.72 | <0.001 |
| PC7 | -1.4 | -2.2, -0.60 | <0.001 |
| PC8 | 1.0 | -0.02, 2.0 | 0.054 |

<sup>1</sup>CI = Confidence Interval

### Pathogen burden associates with higher hs-CRP levels

We then checked if the overall burden of chronic infections contributes to increased hs-CRP levels. Study participants were stratified according to their overall seropositivity index, calculated by summing the number of pathogens for which they were seropositive (range: 0-13). The number of individuals in each serological stratum ranged from 5 (index = 0) to 2’717 (index = 7) and is presented in Fig. 4. We used a linear model to search for an association between pathogen burden and hs-CRP levels. Hs-CRP levels were found to be significantly and positively associated with increasing pathogen burden (P-value = 4.12e-4) (Fig. 4).

**Fig. 4.**
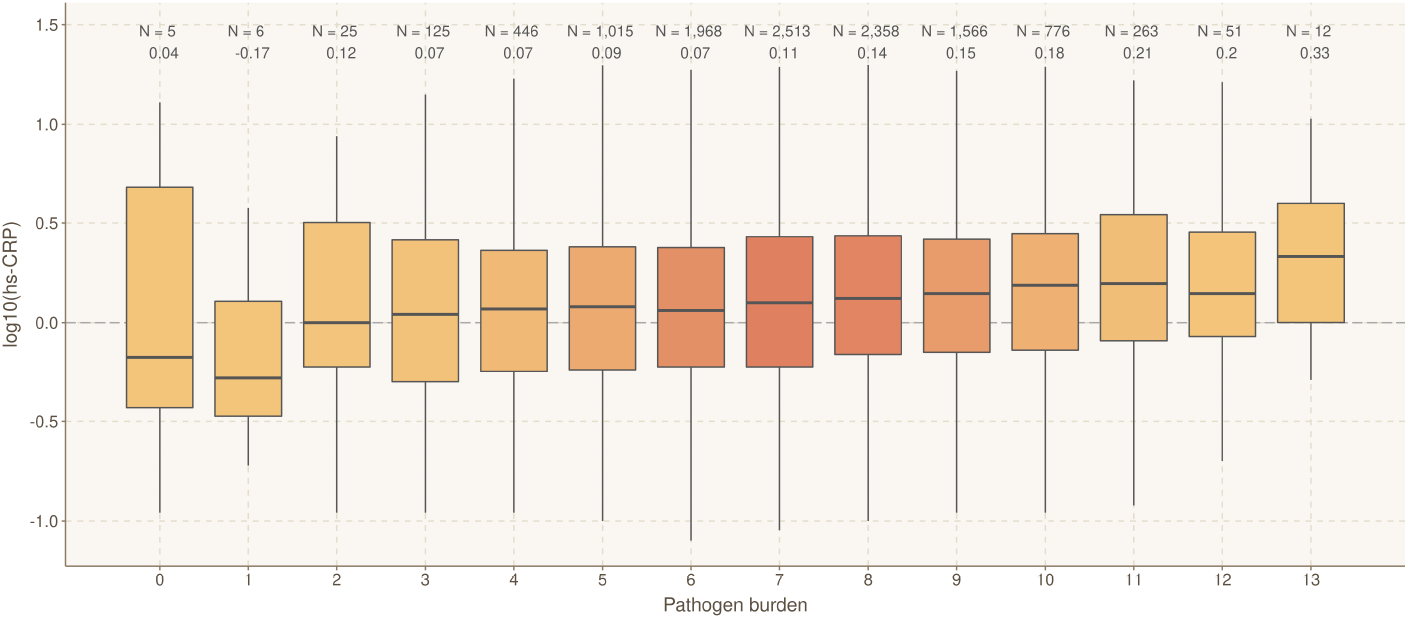
Levels of hs-CRP by infectious burden. Boxplots showing the hs-CRP value for each pathogen load group. The black bold line within the boxplot indicates the median of the hs-CRP measurement. The boxes are colored by sample size. Sample size and median for each group are shown above the box.

## Discussion

Mounting evidence suggests that exposure to multiple pathogens, even when they do not cause obvious disease, can affect the immune system and human health (18, 32, 33). In an effort to better understand the variability of humoral immune response and inflammation patterns in response to pathogen exposure, we selected 27 antigens from 13 persistent infectious agents, which we evaluated using multiplex serology to detect specific immunoglobulin G levels in two well-characterized population-based cohorts.

We first studied the human genetic determinants of hs-CRP levels in our study population. Using a GWAS approach, we confirmed previously reported associations with hs-CRP levels at the *CRP, LEPR, IL6R, GCKR, APOE* and *HNF1A-AS1* loci (9, 28–31). These genes and genomic loci are enriched in hepatic, immune and metabolic pathways. We then investigated the relationship between common genetic variation and hs-CRP levels by calculating a PRS for all study participants. The PRS explained about 4% of the variation in hs-CRP levels, in agreement with previously published results (28). We also found that BMI was the major non-genetic predictor of hs-CRP, with approximately 19% of the variance explained. Next, we studied the impact of persistent infections on chronic inflammation after adjustment for known influencing factors, including age, sex, BMI, and human genetic variability, as explored above. We observed an association between increased levels of hs-CRP and seropositivity for *C. trachomatis* and *H. pylori*. The two gram-negative bacteria *C. trachomatis* and *H. pylori* do not cause life-long, latent infections. Nevertheless, they are responsible for some of the most frequent chronic infections in humans.

*H. pylori* can colonize the gastric epithelium for long periods of time, leading to chronic inflammation of the gastric mucosa. Even if the majority of individuals infected with *H. pylori* have no symptoms, the bacterium has been causally linked with gastritis, gastric ulcer and an increased risk of gastric cancer (34, 35). Our results suggest a systemic impact of chronic *H. pylori* infection beyond the known local inflammatory effect on the gastric mucosa, confirming an observation made previously in a cross-sectional population-based study (36).

*C. trachomatis* causes genital and ocular infection. The ocular manifestation of the infection, trachoma, is the world’s leading cause of preventable blindness and is endemic in many developing countries. This clinical presentation is however highly unlikely to contribute to the 25% seroprevalence of anti-Chlamydia antibodies detected in the Swiss and UK cohorts included in our study. More relevant here, *C. trachomatis* is the etiological agent of human chlamydia urogenital tract infection, which is the most common bacterial sexually transmitted disease. Chronic or recurrent forms of the disease are frequently observed. To our knowledge, no study has examined the direct association between *C. trachomatis* infection and hs-CRP levels at the population level. However, studies conducted in the context of associations between *C. trachomatis* and tubal factor-related subfertility and preterm delivery have also shown elevated hs-CRP levels (37–39).

Altogether these results confirm the role of chronic or recurrent bacterial infections in low-grade inflammation, reflected by a small but consistent increase in hs-CRP levels in seropositive individuals. In addition, we found an association between increased pathogen burden and hs-CRP levels by stratifying individuals according to their cumulative number of positive serological results. This indicates that latent infections might play an enhancing role on chronic low-grade inflammation, even if that effect is too small to be detected at the individual pathogen level.

Previous studies have shown that pro-inflammatory cytokines and chronic inflammation are associated with cellular aging (“inflammaging”) and a number of non-communicable diseases, including certain cancers, type 2 diabetes, and cardiovascular disease (3, 4, 40, 41). It would therefore not be surprising to find that infections also play a key role in these diseases and that the reactivation of these pathogens can contribute to the deterioration of the overall health of older individuals. Finally, CRP-PRS was also found to be significantly associated in the analysis including both genetics and serological results, confirming that human genetic variation plays a modulating role in systemic inflammation.

Our study has some limitations. Firstly, we cannot rule out the effects of other non-measured infections at the time of hs-CRP measurement that may have influenced the level of inflammatory biomarker. Also, we did not adjust our models for all known influencing factors (e.g. smoking, anti-inflammatory or anti-infective drugs, or possible inflammatory diseases). However, participants in both studies were assumed to be in good overall health at the time of data collection, and the data were filtered before analysis to detect levels indicative of acute infection. Secondly, some pathogens had relatively low or high seroprevalences and should be re-examined in a larger study. In particular, it will be interesting to repeat the analysis once serological data for all individuals in the UK Biobank are available. This will allow for greater reliability in terms of statistical power. Third, hs-CRP was the only inflammatory biomarker studied. Other pro-inflammatory cytokines such as IL-1b, IL-6, and TNF-a, are regulators of host responses to infection and positive mediators of inflammation. Consideration of these other biomarkers would give insight into more specific inflammatory pathways and provide a more comprehensive picture of the overall inflammatory status. Fourth, we only observed associations with the presence of chronic inflammation, and our study design does not allow us to infer any kind of causality. In particular, we can’t exclude the possibility that higher levels of inflammation are responsible for the reactivation of a pathogen, resulting in detectable seropositivity.

In conclusion, we found that seropositivity for *C. trachomatis* and *H. pylori* antigens is associated with increased levels of hs-CRP. Together with demographic, clinical and genetic factors, persistent infections contribute to chronic low-grade inflammation, which can have deleterious long-term consequences on health.

## Data Availability

All data produced in the present study are available upon reasonable request to the authors.

## ACKNOWLEDGEMENTS

We thank the participants in the UK Biobank and CoLaus|PsyCoLaus study for their time and contribution to this study. We also thank all the clinical, academic and administrative collaborators who helped with participant recruitment, study coordination, data collection and storage.

## AUTHOR CONTRIBUTIONS

FH: Conceptualization, Methodology, Software, Formal analysis, Visualization, Writing – Original Draft. ON: Formal Analysis. CB: Formal Analysis. NBr: Resources. NBe: Resources. TW: Investigation, Resources. PMV: Resources, Data Curation, Investigation. PV: Investigation. JF: Funding Acquisition, Project administration, Supervision, Writing – Original Draft. All co-authors reviewed the manuscript.

## CONFLICTS OF INTEREST

The authors have no related financial or non-financial interests to disclose.

## FUNDING

This project was supported by the Swiss National Science Foundation (grant 31003A_175603 to JF). The CoLaus|PsyCoLaus study was and is supported by research grants from GlaxoSmithKline, the Faculty of Biology and Medicine of Lausanne, and the Swiss National Science Foundation (grants 3200B0_105993, 3200B0_118308, 33CSCO_122661, 33CS30_139468, 33CS30_148401 and 33CS30_177535/1).

## DATA ACCESS

The CoLaus|PsyCoLaus cohort data used in this study cannot be fully shared as they contain potentially sensitive patient information. As discussed with the competent authority, the Research Ethic Committee of the Canton of Vaud, transferring or directly sharing this data would be a violation of the Swiss legislation aiming to protect the personal rights of participants. Non-identifiable, individual-level data are available for interested researchers, who meet the criteria for access to confidential data sharing, from the CoLaus|PsyCoLaus Datacenter (CHUV, Lausanne, Switzerland). Instructions for gaining access to the CoLaus|PsyCoLaus data used in this study are available at (www.colaus-psycolaus.ch/professionals/how-to-collaborate/).

## Notes

### Competing Interest Statement

The authors have declared no competing interest.

### Author Declarations

Ethical approval for the UK Biobank study was obtained from the North West Centre for Research Ethics Committee (11/NW/0382). The institutional Ethics Committee of the University of Lausanne, which afterward became the Ethics Commission of Canton Vaud (www.cer-vd.ch) approved the baseline CoLaus|PsyCoLaus study (reference 16/03, decisions of 13th January and 10th February 2003), and written consent was obtained from all participants.

