## Supplementary Material for "The combined impact of persistent infections and human genetic variation on C-reactive protein levels"

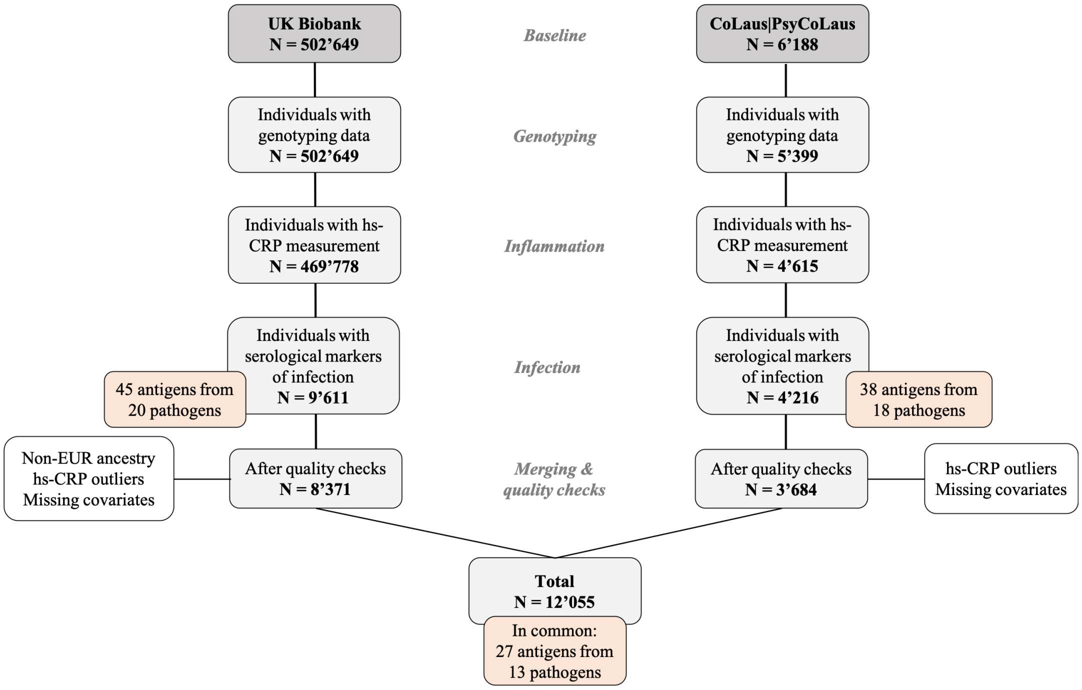


**Supplementary Figure 1. Flowchart illustrating the inclusion/exclusion of individuals in the study.** Orange boxes indicate the number of included antigens and pathogens.

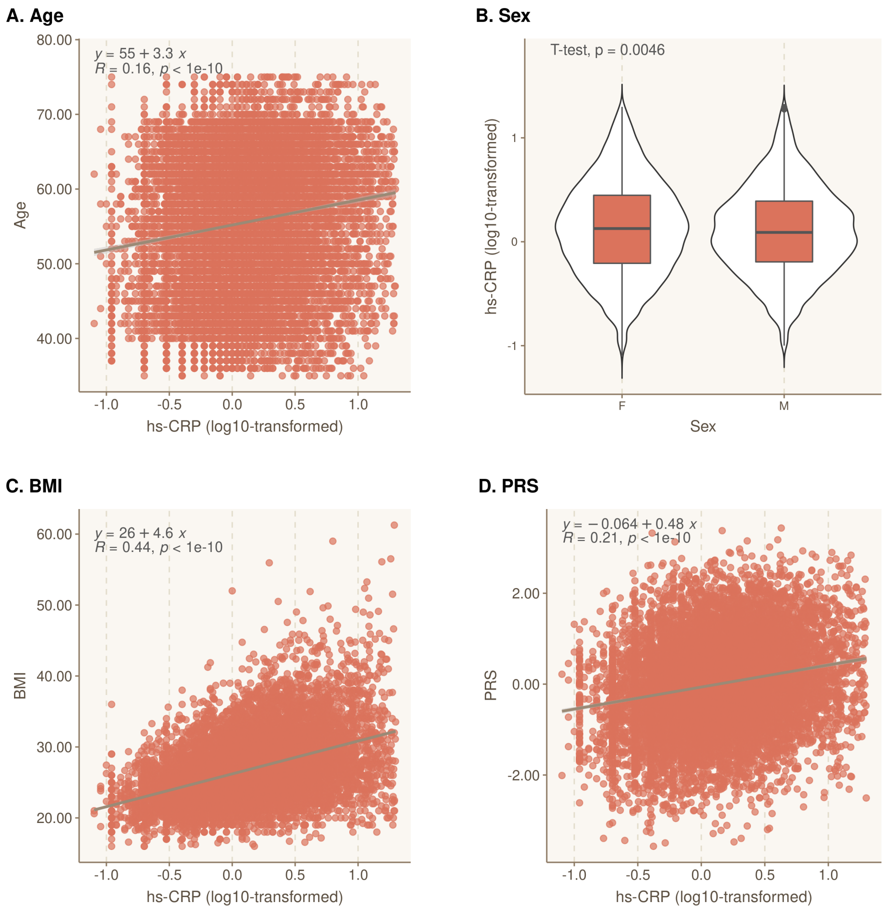


**Supplementary Figure 2. Scatterplot and regression line (with 95% confidence intervals) to describe the relationship of hs-CRP with characteristics of study participants.** Relationship between hs-CRP and A) age, B) sex, C) BMI and D) polygenic risk score (PRS). For linear regressions, linear regression equation, R-squared and P-value are shown.

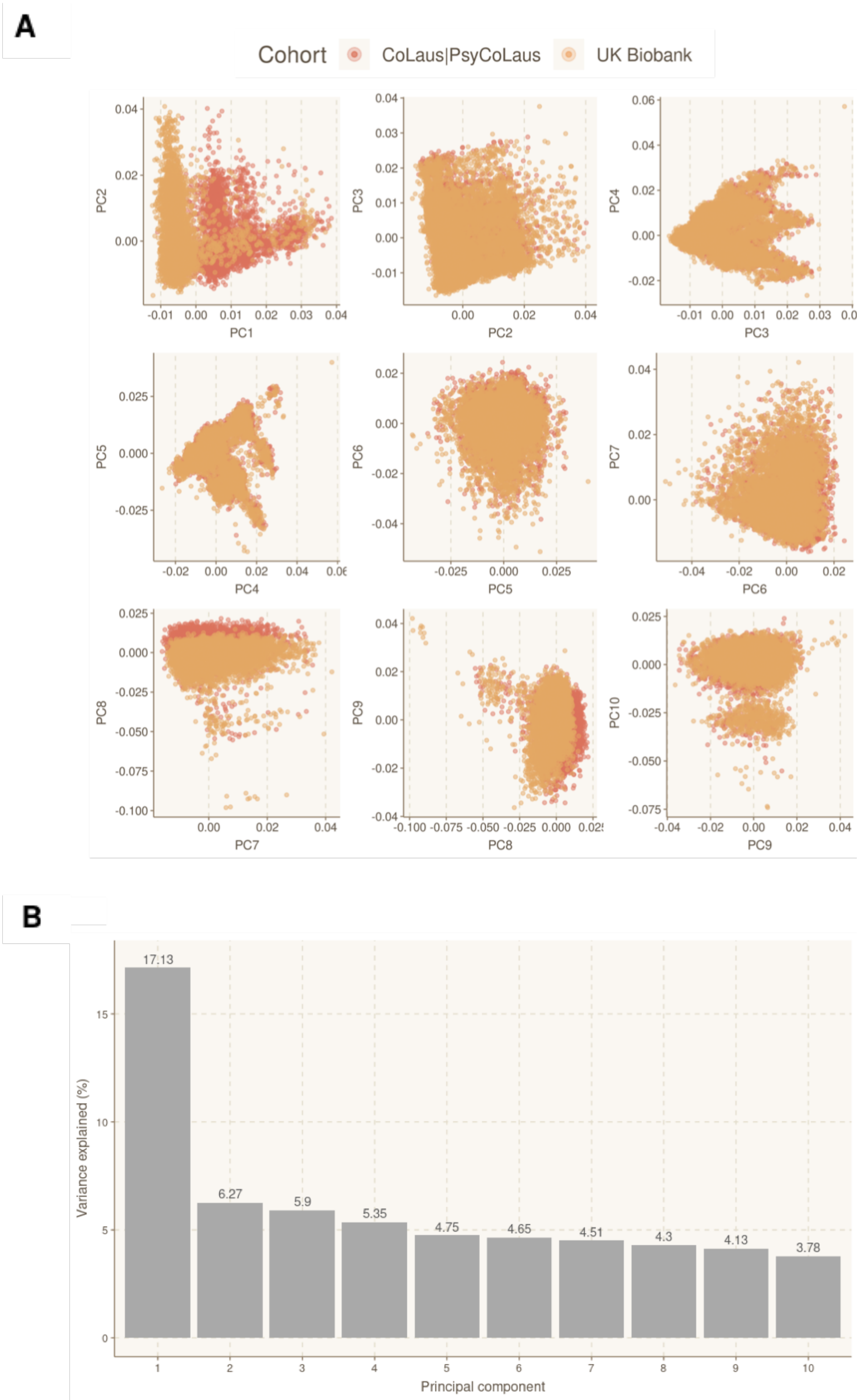


**Supplementary Figure 3. Principal component analysis (PCA) of combined genotyping data.** A) PCA plot of the first ten PCs of the genotyping data. Samples are colored by cohort. C) Histogram explaining the variance of each PC component. In the histogram, the variance explained by each eigenvalue is labeled on top.

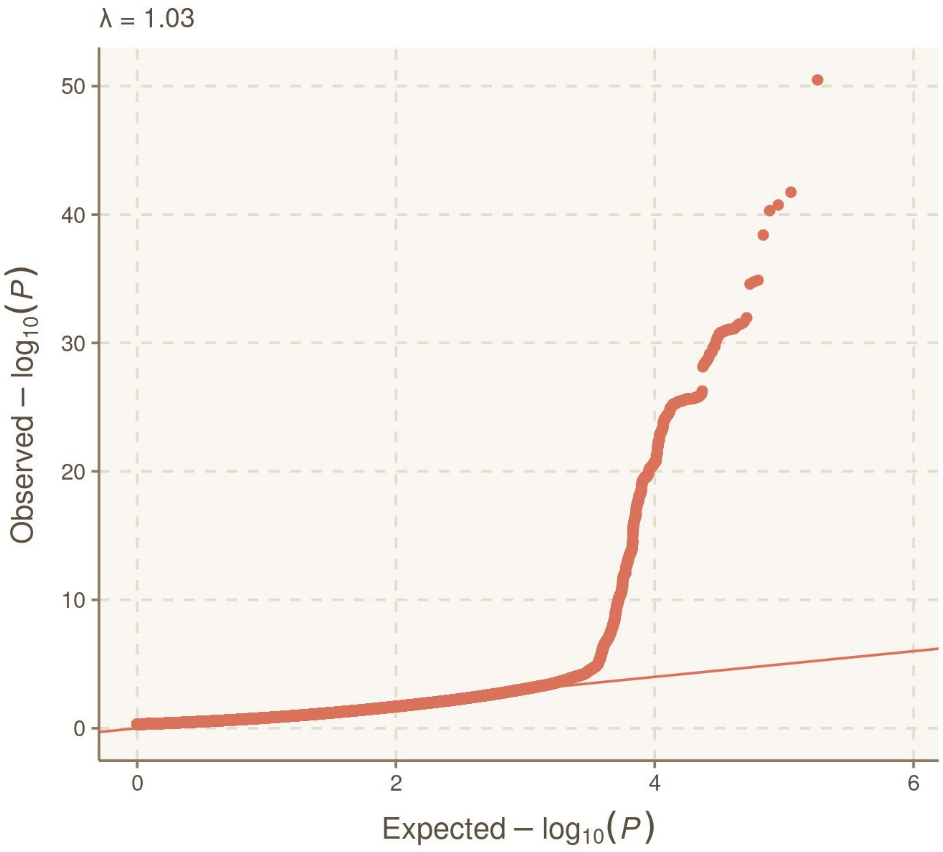


**Supplementary Figure 4. Quantile-quantile (Q-Q) plot of genome-wide association results of hs-CRP levels.** Q-Q plot of SNPs for association with hs-CRP. On the y-axis, the observed P-values (orange dots) are plotted against the expected P-values under the null distribution. The orange line indicates the distribution of SNPs under the null distribution. Lambda (λ) denotes the genomic control inflation factor.

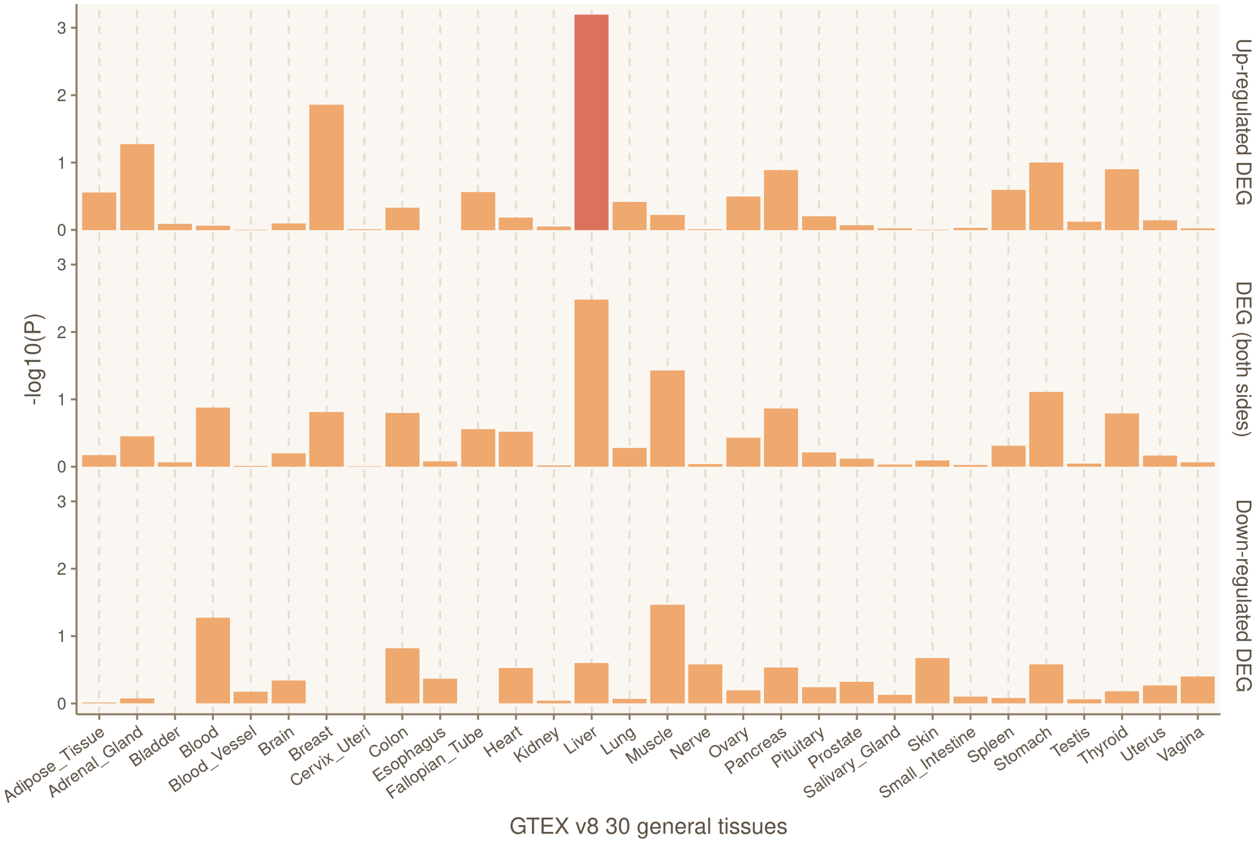


**Supplementary Figure 5. Tissue-specific expression of significant genes relative to hs-CRP levels.** Differentially expressed genes (DEG) plot for hs-CRP levels in 30 general tissue types from GTEx v8. Significantly enriched DEG sets (P-value $\leq$ 0.05) are highlighted in red. Expression values show the average of expression of normal tissues based on GTEx database.

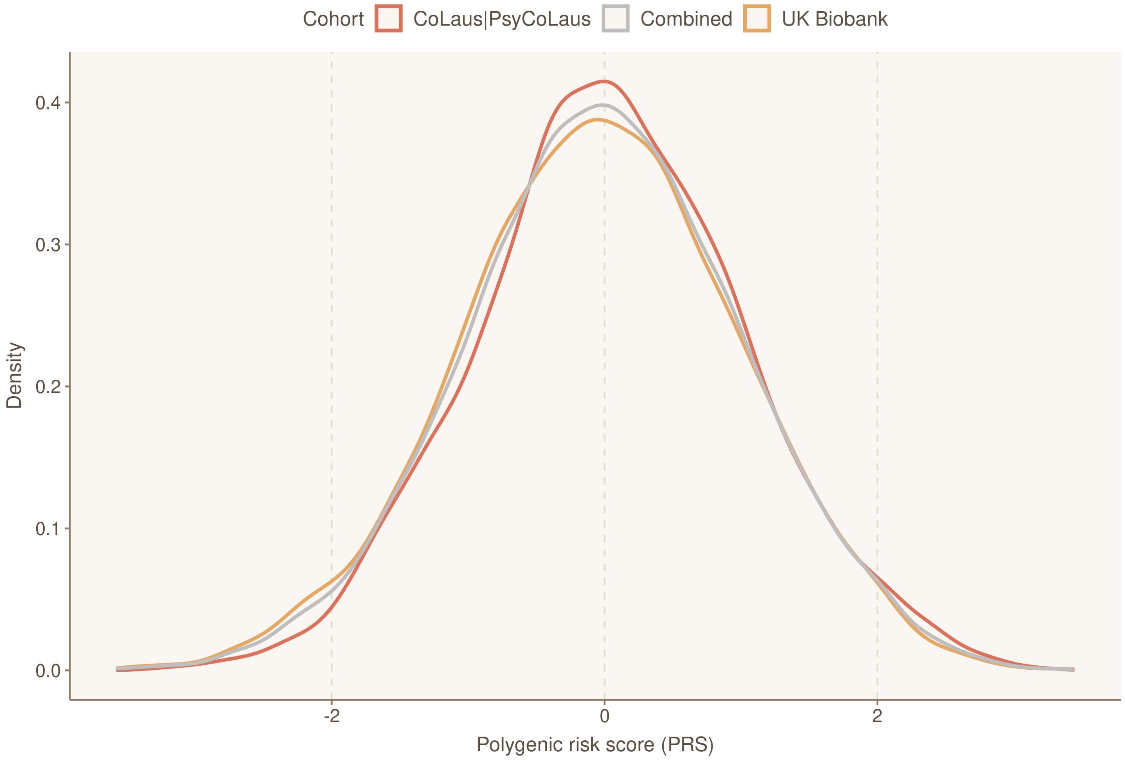


**Supplementary Figure 6. Distribution of polygenic risk score (PRS) values.** Density distribution of standardized PRS values by subcohort (CoLaus|PsyCoLaus and UKB) and across all participants (combined).

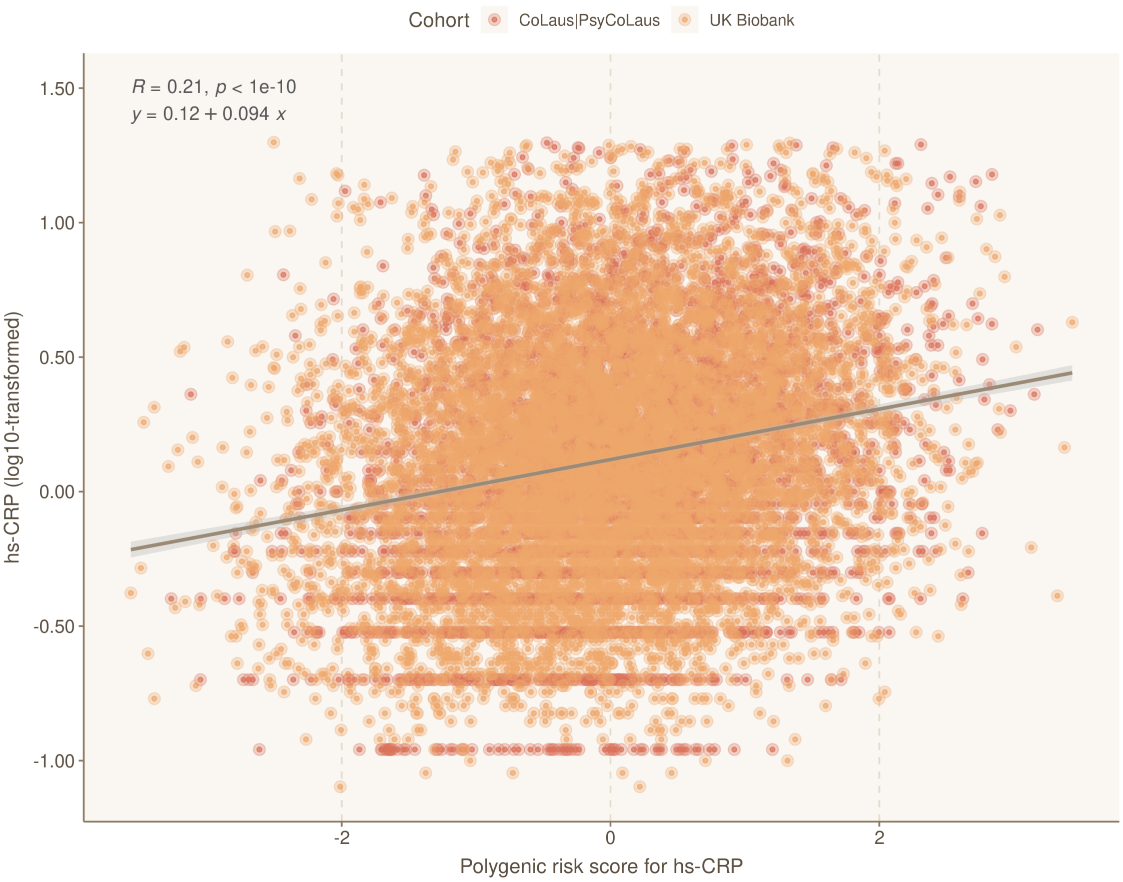


**Supplementary Figure 7. Polygenic risk score for hs-CRP (CRP-PRS) was significantly associated with hs-CRP levels.** Scatter plots with linear regression line of polygenic risk scores predicting hs-CRP levels for individuals in the cohort. 95% confidence interval is showed in grey shade.

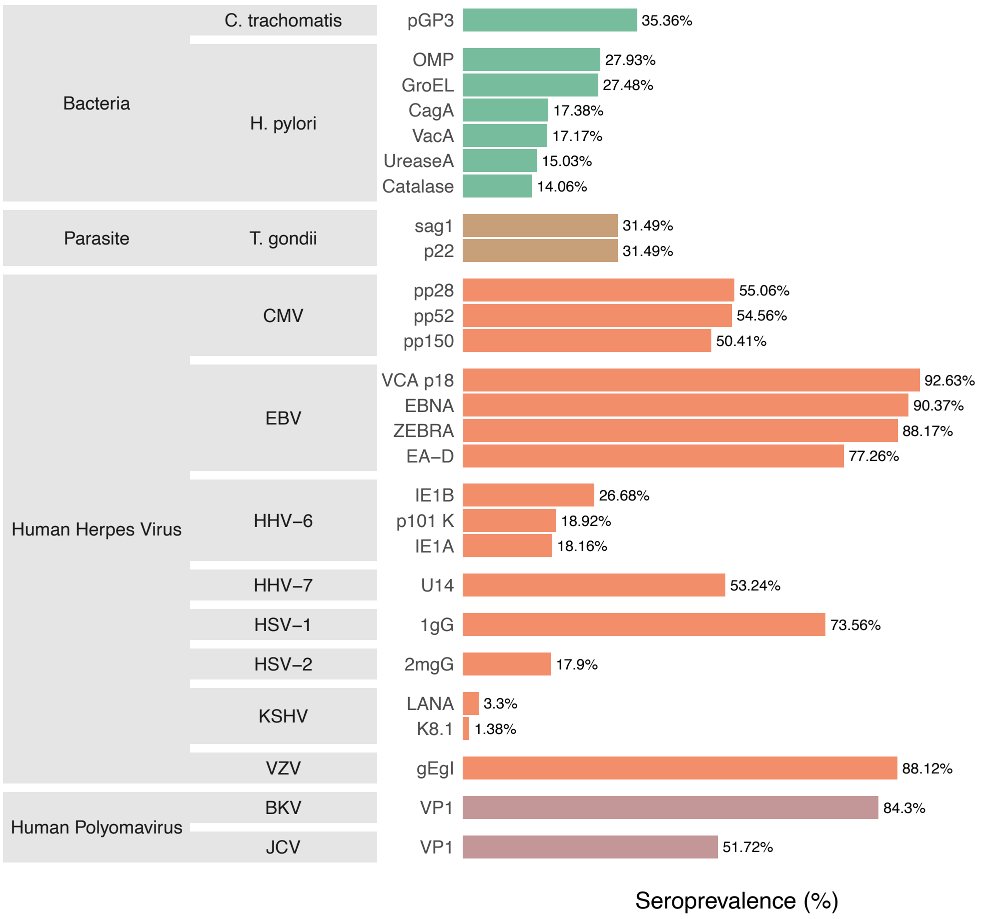


**Supplementary Figure 8. Seroprevalence of tested antigens in the CoLaus|PsyCoLaus.** List of the 27 antigens available from the CoLaus|PsyCoLaus study that are shared with the UK Biobank. The percentages indicate the seroprevalence of antibodies against infectious disease antigens tested using Multiplex Serology platform. The grey boxes indicate the pathogen on which the antigen protein is found, and the family to which the pathogen belongs.

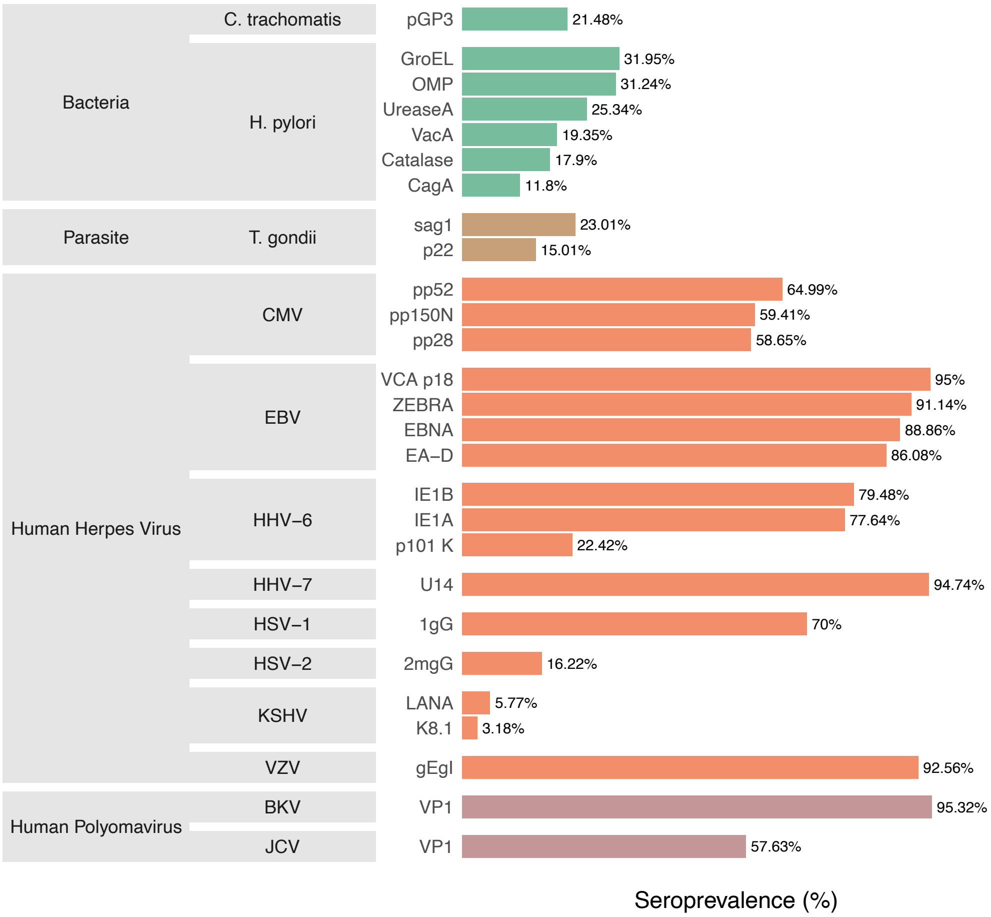


**Supplementary Figure 9. Seroprevalence of tested antigens in the UK Biobank.** List of the 27 antigens available from the UK Biobank that are shared with the CoLaus|PsyCoLaus study. The percentages indicate the seroprevalence of antibodies against infectious disease antigens tested using Multiplex Serology platform. The grey boxes indicate the pathogen on which the antigen protein is found, and the family to which the pathogen belongs.
